# A qualitative framework for understanding hearing-aid gain self-adjustment

**DOI:** 10.64898/2026.07.31.26359402

**Authors:** Janin Benecke, William M. Whitmer

## Abstract

Studies employing and evaluating hearing-aid self-adjustments frequently label the resulting settings as “preferences” without questioning that label. There is a lack of inquiries into the rationale behind participants’ choices or their approach to self-adjustment. We here employed a think-aloud protocol concurrent with individuals’ self-adjustments to capture qualitative perspectives on these processes. Thirty-six adult hearing-aid users were asked to verbalise what they were thinking while adjusting to their preference three stimuli (music, speech or speech in noise) using three control interfaces (1 slider jointly controlling bass/treble, 2 sliders separately controlling bass and treble or 3 sliders controlling bass, mid and treble). Audio-video recordings were transcribed, annotated and analysed using inductive content coding and thematic analysis. Participants’ approach and navigation of the self-adjustment process was best defined as a continuum between exploratory and anticipatory archetypes, with both types sometimes occurring at different stages within the same adjustment. The exploratory type initially assesses control functions, then selects the optimal setting from available options, and evaluates sound changes affectively in isolation or comparatively against prior settings. The anticipatory type begins by evaluating the sound, identifying issues, adjusting settings guided by past experiences, and evaluating adjustments against the identified issues. Additionally, the descriptors elicited during adjustments generally agreed with expert-based data. The resulting qualitative framework of self-adjustment helps explain both how hearing-aid users navigate the personalisation process and how self-adjustment can promote greater understanding of the options available and greater ownership in that process.

## INTRODUCTION

People with hearing loss differ widely in their supra-threshold hearing abilities (e.g., Bentler & Cooley, 2001; Füllgrabe & Moore, 2018), everyday listening environments (e.g., Gatehouse et al., 2006), needs (e.g., Pryce et al., 2026) and sound-quality preferences (e.g., Vaisberg et al. 2021). Frequency dependent gains, the fundamental point of personalisation in hearing aids, are prescribed to fit an assumed *average* listener with given audiometric pure-tone thresholds, often interpolated from a small number of participants. Consequently, prescriptions have long been regarded as only starting points for further personalisation to better meet individual needs (e.g., Kuk, 1999; Schweitzer et al., 1999; Keidser et al., 2012; Kollmeier & Kiessling, 2018). Standard clinical practice for personalisation is to adjust hearing-aid settings (e.g., louder/softer for lower/higher-pitched sounds) based on responses to the clinician’s voice (e.g., ‘*How does that sound?*’). Clinical personalisation practice, however, has theoretical and psychometric flaws: clinicians cannot perceive sound as patients do, changes may not be immediately perceptible nor verifiable to the hearing-aid user (Caswell-Midwinter & Whitmer, 2019; Whitmer et al., 2022) and patients’ descriptions of what they hear can be unreliable and inconsistent (Caswell-Midwinter & Whitmer, 2021). Additionally, recalling and reflecting on acoustically complex listening situations one has experienced can be difficult in the clinic, where the clinician’s and patient’s voices are often the only readily available stimuli. Given these issues, hearing-aid gain personalisation in the clinic risks becoming a futile, frustrating and protracted procedure (Cunningham et al., 2001). An alternative approach is to involve the hearing-aid user directly and actively in the personalisation process by letting them adjust hearing-aid gain parameters themselves. This approach has been implemented in the burgeoning number of over-the-counter hearing aids that users fit themselves without the help of a clinician, as well as in apps for clinically fit and over-the-counter hearing aids that control settings via smartphones (e.g., Sabin et al., 2020). In any of these cases, user manipulation of frequency-gain parameters requires mapping these parameters to controls that are easy to use and understand and perceptually relevant and achieve their myriad aims and preferences.

Self-adjustment methods differ in how parameters are combined and presented in user interfaces. For some methods, controls are labelled by function. Examples include the *Goldilocks* interface (Boothroyd & Mackersie, 2017; Mackersie et al. 2020; Boothroyd et al. 2022), which labelled up/down buttons ‘loudness,’ ‘crispness’ and ‘fullness’ to adjust overall gain, high-frequency and low-frequency tilt, respectively, and the *Ear Machine* interface (Nelson et al., 2018; Perry et al., 2019; Sabin et al., 2020) which labelled two wheels ‘loudness’ and ‘finetuning’ to control overall gain and tilt, respectively. Other methods have used different visual representations to avoid functional labels, such as settings represented as points on a 2dimensional parameter space (e.g. Goβwein et al., 2023; 2024; Kur§un et al. 2025), or the parameters space is presented as tiles that can be marked (Valentine et al., 2011; see also Debevc et al., 2021), or settings were represented radially on a colour wheel (Yang et al., 2017). Conversely, the PAS (Parameter Adjustment and Selection) method (Punch & Robb, 1992) deliberately avoided labels and visual feedback to attempt to ensure that adjustments were guided solely by acoustics. The diversity in approaches highlights how designing effective hearing-aid self-adjustment interfaces is as much a human-computer interaction (HCI) problem as a psychoacoustical one. Labels can provide clarity and guide use, but they may confuse participants unfamiliar with the terminology, induce expectations and/or risk influencing participants to interact with controls according to what they read rather than what they hear. Additionally, Caswell-Midwinter and Whitmer (2021) found very little consistency in the application of sound descriptors to changes in frequency-gain response (FGR). Conversely, label-less or ‘blinded’ methods can shift the focus towards auditory perception but may reduce transparency and learnability. Despite the question of how best to provide FGR self-adjustment being an HCI problem, it has rarely been treated as such. In the current study, we apply a common HCI technique (Nielsen, 1992) to explore self-adjustment useability from a qualitative perspective.

A key component in assessing self-adjustment useability is understanding what individuals intend to achieve; that is, their criteria in navigating FGR self-adjustment may affect their chosen FGR. Such internal criteria may be shaped by external factors. Keidser et al. (2005) and Goβwein et al.(2023), for example, instructed participants (i.e., provided external criteria) to optimise FGR for speech understanding and comfort, resulting in generally more gain for the speech-understanding criterion. Although the impetus for imposing external criteria is to reduce variability, there still were substantial interindividual variances in these studies, possibly indicating differences in how participants interpret or prioritise these criteria. Not using external adjustment criteria may elicit more inherent, ‘natural’ self-adjustment behaviours at the cost of increasing variance. The relative importance of individual criteria may also depend on the initial conditions, including the type of stimulus. For example, if clarity is already adequate at the start of an adjustment, it may carry less weight as a criterion than when clarity is poor (cf. McShefferty et al., 2016). Numerous self-adjustment studies have shown differences in adjusted FGRs due to the presence, type or level of noise (e.g., Keidser et al., 2005; Nelson et al., 2018; Goβwein et al., 2023). Mackersie et al.(2020)found that after *Goldilocks* FGR self-adjustment without a priori criteria setting, participants predominantly chose *loudness* and *clarity* as equally important for speech in quiet, but only *clarity* for speech in noise. In a subsequent Goldilocks study, participants’ most important criterion varied more between *loudness*, *quality*, *intelligibility* and *comfort*, and crucially, participants reported difficulty in selecting a single criterion, suggesting they balance multiple criteria and seek compromises during adjustments (Boothroyd et al. 2022). Overall sound impression is generally acknowledged as being multi-dimensional (e.g., Gabrielsson, 1979; Pedersen, 2015), hence it is not surprising that the actual criteria used by participants in FGR self-adjustment are likely complex and dynamic. A critique of self-adjustment is that, in the absence of a clear perceptual understanding or clearly defined adjustment goals, individuals may rely on trial-and-error strategies, which may increase the likelihood of unsuccessful outcomes (Jensen et al., 2019). Such trial-and-error adjustments, however, may help individuals define their goals. The goal in self-adjustment is subjective and potentially evolving through the process. It is therefore important to evaluate whether self-adjustment is experienced as challenging and to pinpoint the nature of these challenges.

Previous analyses of self-adjustment behaviour have mostly been limited to duration or button presses (e.g., Dreschler et al. 2008)as a proxy for task efficiency. Goβwein et al.(2024) analysed the movements made across a 2-D surface that controlled overall gain on one axis and spectral tilt on the other. Based on this quantitative data, they found four clusters of interface interaction behaviour that were not related to individuals’ adjusted FGR: *curious*, *cautious*, *semi-browsing*, and *full-on browsing*. Furthermore, studies involving the 2-D surface interface (Kliesch et al., 2023; Gδβwein et al., 2024)found that exploration was often aligned with the interface’s axes (i.e., adjusting gain then centre frequency or vice versa). Exploration of a single dimension could have been a strategy to simplify the two-dimensional parameter space to better understand its function/perceptual effects. Because experiences during self-adjustment are subjective and can involve complex trade-offs, the hearing-aid user’s experience can be difficult to capture accurately retrospectively.

Understanding the relation between self-adjustment aims, behaviours and outcomes may not be possible without understanding what motivated the differences in behaviour, or more specifically, accounting for the individuals’ intentions, their approaches to self-adjustment, whether compromises were made, and which criteria guided adjustment decisions. Such understanding can be gained through the think-aloud protocol (TAP), a qualitative method largely attributed to Ericsson & Simon (1980; 1984; 1993) but rooted in the practice of introspection (*Selbstbeobachtung*, e.g., Wundt, 1888) and most associated with usability or user-experience testing (e.g., Nielsen, 1992; Rosenzweig, 2015), although it has been used in many fields (Güss, 2018). While introspection has long been critiqued in behaviourism as interfering with thought processes (e.g., Watson, 1925; Skinner, 1977) or lacking direct access to those processes (Nisbett and Wilson, 1977), but this limitation primarily applies to processes that never reach conscious awareness (Wilson, 1994). The focus of TAP studies is not on whether a task is solved correctly, but rather on understanding the cognitive processes involved (Newell & Simon, 1972). In the literature, the term TAP has been used both to refer to the result (i.e., “protocol” as in the verbalised thought pattern) and to the method (i.e., “protocol” as in procedure). Here, we refer to TAP as a method. TAP is designed to gain insights into participants’ in-the-moment thought processes (van Someren et al., 1994) by recording and analysing verbal protocols of participants’ verbalised thoughts. In hearing research, TAP has been used to develop hearing-related web tools to provide assistance to new hearing-aid users (Maidment et al., 2020) and help-seeking (Hickson et al., 2024), but not, to our knowledge, in the context of self-adjustments. We here employed concurrent TAP to investigate (1) how do participants approach and navigate self-adjustment, (2) what drives decision making during self-adjustment, and (3) how are changes in frequency-gain experienced both generally and as a function of interface controls and stimuli.

## METHODS

Participants had prior exposure to interfaces and stimuli by first completing two related tasks: self-adjustment to preference in a previous session and sound-matching to endpoints earlier in the same session (Benecke & Whitmer, 2026). During TAP, some participants commented on this familiarity, particularly in reference to the stimuli, but also the interfaces.

### Participants

TAPs were completed by 36 adult participants (13 female), aged 61-82 (median 72.5). Participants were recruited via postal invitation from the Hearing Sciences 一 Scottish Section participant pool based on the criteria of having mild-to-moderate losses and being bilateral hearing aid users. Pure-tone thresholds were measured 29-153 days previous to TAP and are shown in the left panel of Figure 1. Better-ear four-frequency (0.5, 1, 2, and 4 kHz) pure-tone threshold averages (BE4FAs) ranged from 18.75-56.25 dB HL (median 38.75 dB HL). Ages and BE4FAs were not correlated (*r* = −0.10; *p*》0.05). Participants were all current bilateral hearing-aid users with 1-28 years (median 7 years) of hearing-aid experience. From each participant’s better-ear audiogram, NAL-R gain prescriptions (Byrne & Dillon, 1986) were calculated to serve as the starting gain for adjustments on each trial (right panel of Figure 1). These individualised but antiquated and unfamiliar gains were used to promote self-adjustment away from the initial baseline.

**Figure 1.**
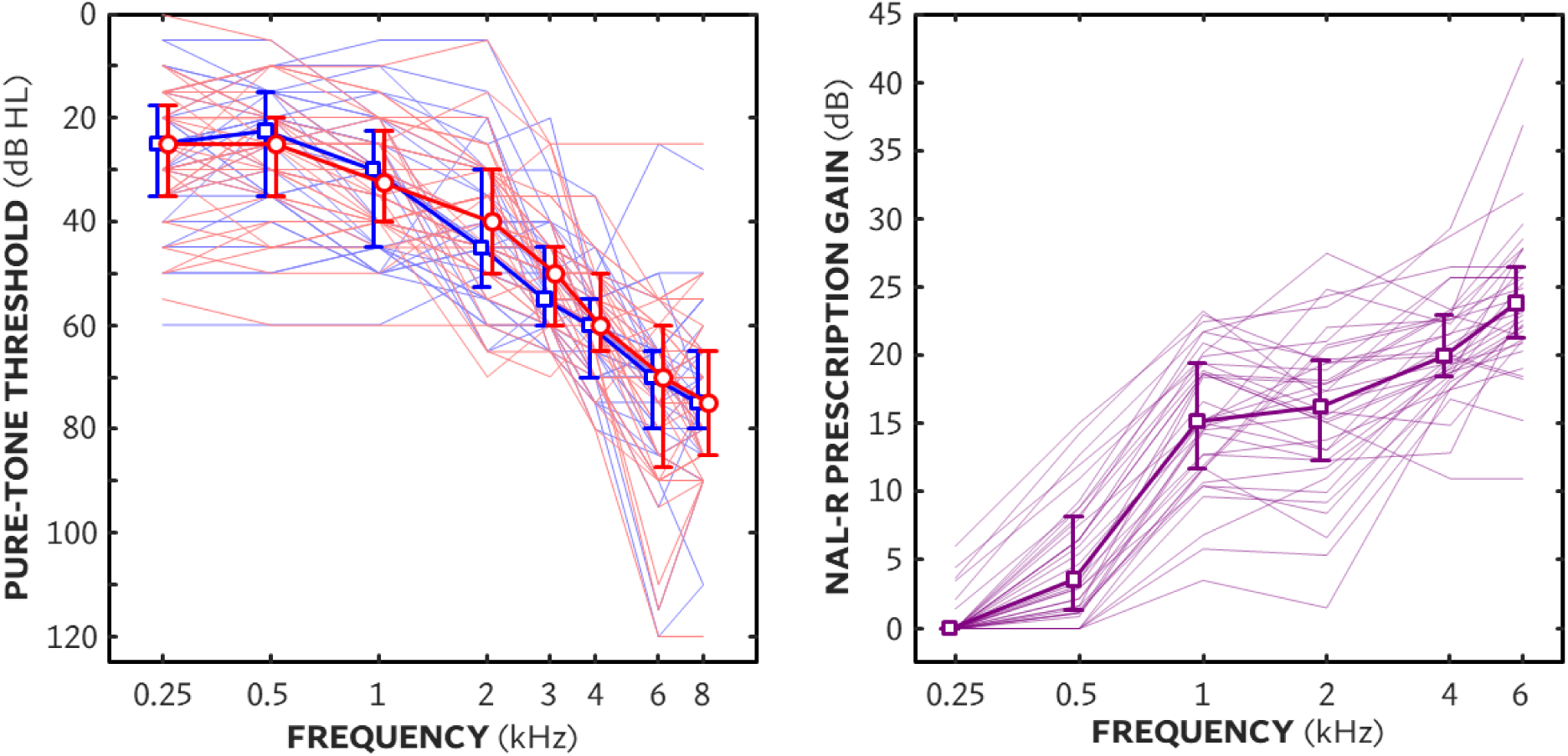
Left panel shows hearing thresholds as a function of pure-tone frequency for left (blue) and right (red) ears. Right panel shows NAL-R prescription gain - the initial gain on each trial - as a function of frequency. Thin lines show individual audiograms/gain prescriptions. Symbols and error bars show medians and interquartile ranges, respectively. Right panel shows prescription formula (NAL-R) gains, used as the starting point for each adjustment, as a function of frequency.

All participants gave written informed consent prior to taking part and were paid a small allowance for their participation. Participants were specifically informed that the use of the audio-visual recordings was strictly limited to create the protocols (transcriptions and annotations of audio-visual material). All procedures were approved prior to recruitment by the North East - Newcastle & North Tyneside 2 Research Ethics Committee (23/NE/0071).

### Apparatus and stimuli

Participants sat at a desk with the touchscreen inside a double-walled audiometric booth (IAC Acoustics). Participants adjusted the FGR to preference from an initial individual NAL-R prescription (right panel of Figure 1) for three different stimulus types (speech in cafeteria noise, speech in quiet and music) using three different interfaces with 1, 2 or 3 unlabelled vertical sliders controlling ±18 dB gain across different frequency bands as shown in Figure 2. Sliders controlled the gain of 2nd-order IIR filters with coefficients set to produce a flat response (±1 dB in third-octave bands from 100-8000 Hz) when the sliders were increased or decreased by the same amount. Slider resolution (i.e., the smallest change in gain possible) was 0.036 dB. The speech (in-quiet) stimuli consisted of segments a male talker with central Scottish accent reading *The Musgrave Rital* by Sir Arthur Conan Doyle (Macpherson & Akeroyd, 2013). The speech-in-noise stimuli consisted of different segments from the same speech recording in cafeteria babble noise (Bjerg & Larsen 2006) at +5 dB SNR RMS. The music stimuli were segments from Barry White’s “I Can’t Get Enough Of Your Love Babe” (White, 1974). Three unique segments for each stimulus type 一 one for each interface 一 were used with durations ranging from 14-21 s (mean 17 s) to maintain the varying ends of speech and musical phrases. All stimuli were gated with 25-ms raised-cosine onset and offset ramps.

**Figure 2.**
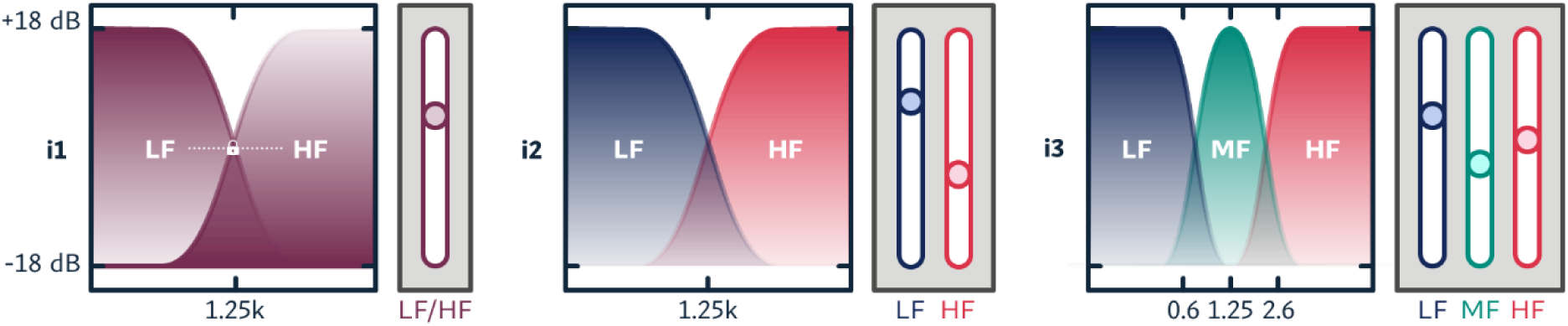
Schematic of filter functions for the three interfaces illustrating the ±18-dB adjustable range as a function of frequency for each interface. Left panels show interface 1 (i1), a single slider adjusting LF and HF gain in opposite directions. Middle panels show interface 2 (i2), two independent sliders for LF and HF. Right panels show interface 3 (i3), three independent sliders for LF, MF and HF.

Participants’ verbalisations and behaviours were recorded using a Yeti Nano tabletop microphone (Logitech International S.A., Lausanne, Switzerland) for audio, and an ELP 4K USB camera (Shenzhen Ailipu Technology Co., Ltd., China) for video. Both were recorded synchronously using the Windows Camera app (version 2023, Microsoft Corporation, Redmond, USA) on an independent desktop PC not running the experiment. For simplicity, the combined audio and video recordings are hereafter referred to as videos.

### Procedure

All procedures including TAP were conducted by the first author and pilot-tested with four members of the Hearing Sciences 一 Scottish Section Patient and Public Involvement Group. The task and procedure were explained to participants verbally and available in writing throughout the experiment, and participants were encouraged to ask questions. Instructions emphasised that there were no wrong or right adjustments, and that they may be reminded by the experimenter to keep talking if they were silent for too long (specifically in relation to interactions that may have been motivated by a specific intention; e.g., switching from one slider to the next).

#### Think-aloud practice: Image brightness adjustment

Participants first completed four practice TAP trials prior to experimental trials. The purpose of these practice trials was to help participants to become familiar with verbalising their thoughts, such as recognizing the faster pace of thinking compared to speaking or adapting to prompts reminding them to keep talking if they remained silent for extended periods. While Ericsson & Simon (1984) employed ‘warm-up’ trials in problem solving, such as asking participants to think aloud while solving an arithmetic problem, van Someren et al. (1994) recommended to choose a practice task similar to but still unrelated to the target task to not bias participants’ initial thoughts. The TAP practice trials involved adjusting image brightness to preference (‘*Please adjust the image to your preference. While doing so, say out loud anything that goes through your mind.*’) using a single slider on the same touchscreen used with the self-adjustment interfaces. Royalty-free images of local (Glasgow) landmarks were used as to be likely familiar to participants and evoke personal memories, which would help initiate verbalisation. On each of the four practice trials, the starting point of the image brightness was randomly offset from the original.

#### Think-aloud: self-adjustment to preference

After the practice session, participants then completed nine self-adjustment-to-preference trials while thinking aloud. Each trial presented a unique combination of stimulus type and interface, with the order randomised across participants. Before starting the recorded trials, participants were reminded that the experimenter could not hear what they heard, and that their task was to verbalise (‘say out loud’) their thoughts while adjusting to preference. They were also reminded that they could pause the audio at any time with the corresponding button. If participants fell silent during the adjustments, in particular when directing their attention to another slider, they were prompted both aurally and visually to continue talking. After the trials, participants had the opportunity to give feedback on their experience performing TAP during self-adjustment.

### Data Analysis

#### Data preparation

All videos were transcribed and annotated by the first author using a style guide developed prior to transcription (see Supplemental Materials). Annotations included interaction with the sliders or interface (e.g., ‘i2HF down’, ‘pauses playback’) and non-verbal communication (gestures, facial expressions (e.g., ‘points at i2HF’, ‘hand gesture opening and closing a mouth; talking’). Transcripts also included prompts of the experimenter (e.g., [E: please keep talking]). Transcription and annotation were reviewed for completeness and agreement with the videos by the first author and then a random sample was cross-checked independently by the co-author (cf. Whitty et al., 2014). Transcripts were produced in table form, allowing one row per slider interaction, which detailed the minimum and maximum points reached in dB as difference relative to the NAL-R baseline. Additionally, time-series data was plotted for each trial for annotation reference. Transcription and annotation allowed extensive familiarisation with the data, including noting initial impressions and ideas on emerging concepts. Transcripts (of all nine trial per person, that is 324 trials in total) and were imported into a computer-assisted qualitative data analysis software, NVivo version 14 (Lumivero, Denver, Colorado).

#### Reflexive thematic analysis

Think-aloud protocols were assessed thematically to uncover how participants approached self-adjustment and to identify decision-making processes and reactions to differences in FGR. In the absence of an established theoretical framework for self-adjustment, we adopted an inductive approach to qualitative analysis, allowing findings to be grounded in the protocols rather than from a predefined theory. Furthermore, the approach to analysis reflected a deliberate departure from quantitative or post-positivist approaches which assume that meaning can be reliably segmented and quantified across instances and employ procedures intended to mitigate researcher bias, such as consensus coding or the calculation of intercoder agreement and subsequent present theme-frequency counts (Braun & Clarke, 2022). In-line with the observation that the data did not constitute complete or fully articulated verbal reports, our approach treated qualitative analysis as an inherently interpretative practice. Consequently, reflexive thematic analysis (Braun & Clarke, 2006, 2021) was chosen over alternative approaches commonly associated with TAP, such as protocol analysis (Ericsson & Simon, 1993) or line-by-line content analysis (Maykut & Morehouse, 1994).

The six stages from Braun and Clarke (2006) were used as a conceptual framework to guide the inductive analysis: (1) familiarisation, (2) initial coding, (3) theme development, (4) theme review and refinement, (5) theme definition, (6) reporting. This conceptual framework is not a strictly linear process; the stages dealing with the generation and development of initial codes, reviewing and organising codes into themes and the revision of themes form an iterative process (Braun & Clarke, 2006). Meaningful passages that were relevant to performing the adjustment were identified and coded, initially for the transcripts of all trials of ten participants. A verbatim approach was used to generate initial codes, preserving participants’ own words (Saldaña, 2016). This generated a large number of codes without saturation, that is, without reaching a point in which no new codes were required as no new relevant information was coded. Upon coding approximately half of the data and grouping codes by overarching concepts or situations, as well as revising these initial themes, two main themes were identified, both including rich and diverse contents: (1) approaches throughout the adjustment process, and (2) perceptual changes and reactions when adjusting sliders. During theme development, the codes associated with Theme 1 were further organised into three sequential stages (before, during and after the adjustment) that reflected the temporal structure of the adjustment process rather than a collection of stand-alone themes or sub-themes situated within the process. Theme 2 was situated sequentially within the during-adjustment stage of the resultant complete model, Theme 1.

## RESULTS

For all TAP examples of themes and subthemes, pseudonymised participant identifiers are used (e.g., “p10”). As with any interview study, the amount of information 一 the time spent talking 一 varied considerably across participants. All participants provided some thoughts, but there were rare instances of trials without verbalisation, often due to the participant moving on to the next trial just before being reminded to keep talking. Some participants appeared to struggle with sharing their inner thoughts, possibly because it involved revealing their internal stream of consciousness, which may be experienced as inappropriate or uncomfortable (Güss, 2018). TAP results from 27 of the 36 participants are provided in the following sections to exemplify the themes.

### Theme 1: Exploratory - anticipatory spectrum

Organising codes into themes, and iteratively refining those themes (Braun & Clarke, 2006; 2021), formed the foundation of model development in this analysis. When reflecting on the underlying structure and how to organise codes, the analytic approach to the qualitative data shifted from traditional thematic analysis, describing self-contained themes, to a way of preserving the procedural context, that is phases or stages in adjustment, in which they occur. When organising the data, verbalisations were categorised into three macro stages: before adjustment (i.e., before interaction with a slider), during adjustment and after adjustment. Reviewing the data in this way revealed that there is a spectrum of approaches to self-adjustment, guided by what could be described as two archetypes: exploratory and anticipatory.

The exploratory archetype is characterized by an approach to self-adjustment driven by the question: *’What are the best possible settings given the available options?’* In contrast, the anticipatory archetype is guided by the question: *‘How can I achieve the sound that I want?’* The two archetypes or mindsets can be understood as opposite ends of a spectrum, in which mixed forms and crossovers exist, they are however helpful in understanding the different mindsets influencing how participants navigate their own adjustments. The full qualitative model of exploratory-anticipatory behaviours in self-adjustment is shown in **Error! Reference source n ot found.**. The figure shows the possible flow (arrows) of self-adjustment, the possible actions (rounded boxes) and types of thoughts at the left and right, indicative of exploratory and anticipatory archetypes, respectively, for each stage (before, during and after adjustment). Each potential process is discussed by stage in the following sections. A verbose version of the model with exemplar quotes instead of themes is provided in the Supplemental Materials.

#### Before the adjustment

The exploratory approach is typified by initial interactions that are motivated by a desire to understand the function of each control. Comments on the initial sound were typically absent, possibly because the focus was on exploring controls, and hence the sound was expected to change. Characteristic adjustments involved both increases and decreases from the initial mid-point slider position, aimed at gaining perceptual certainty about the effects of each control.

> *Let’s see what this one’s doing. [i2LF up]* (p06)

> *Alright, I’m gonna try the first one and see what happens. [i2LF up]* (p03)

> *This one (i3HF) now. [i3HF up] [mumbles: Not sure] what it does. I go the opposite way and come back down again [i3HF down].* (p13)

The anticipatory approach is characterised by first evaluating the initial sound to identify issues. Verbalisations could include referring to particular aspects or overall impression of the sound (e.g., loudness or spectral balance), and the intention to adjust using the controls in a certain way to improve or resolve issues. To this end, anticipation was guided by prior exposure or experiences with self-adjustment, in particular when participants expressed that the initial sound had consistently the same issues.

> *Oh, it starts off very tinny.* (p16)

> *Oh, right. So, my initial reaction: much too shrill. I prefer a bit of bass in that.* (p08)

> *Straight away, it’s no--for me, it’s not loud enough. The treble is too much, so I go looking for…[i3LF up]…a bit of bass.* (p20)

The anticipatory approach was essentially complaint-driven. When the initial sound was perceived as acceptable, and hence no adjustment goals could be expressed, a crossover to a more exploratory mindset occurred, given that slider interaction was required in each trial.

> *That sounds actually quite nice the way it is. (laughs) So…I think I’m not going to do much. [i3LF down]* (p05)

> *Well, that’s sounding as I expect it to be. It’s…--the sound is as I would have it. But I will try to take it up. [i2LF up, down] Noticed…It’s a bit better when I take that up a bit.* (p10)

#### During adjustment

The exploratory approach during slider interactions tended to test both directions from the initial central slider position. In complex interfaces (i2/i3), adjustments were often made sequentially from the leftmost to the rightmost slider. By manipulating each slider, participants identified the resulting changes, which in turn directed their attention during evaluation. Exploratory evaluations were typically hedonic or affective in nature and were often made relative to the previous setting (e.g., better/worse).

> *Haha. Now, [i2LF down, up, down, up] Bass boost in that one.* (p15)

> *[i2HF up] No. [i2HF down] That’s a bit better.* (p02)

> *[i1 up] Oh, that’s worse, that’s terrible!* (p17)

> *[i3HF up, down] And a bit of treble…-- yeah, I’m quite happy with that.* (p08)

The comparative evaluations of the exploratory approach behaviour can be seen as engagement in selecting an optimal setting from the available options. Crossing over to an anticipatory approach occurred with experience or when an issue was identified or anticipation of a certain outcome occurred.

> *[i3HF down, then all the way up]…I was just playing with the treble. Increasing the treble makes the sibilance very unpleasant indeed.* (p09)

> *Alright [i1 up]…I think 一 see fit’s tinny -- it sounds tinny-1 don’t like that. [i1 down] I like that. It’s more of a deeper, a more comfortable sound. That would do.* (p05)

The anticipatory approach during slider interactions relied on prior experiences and knowledge from prior adjustments to predict or anticipate how each slider affected the sound and how to operate them to achieve a defined adjustment goal. Anticipatory adjustments can be characterised as finding optima within an anticipated range.

> *Just a wee bit treble-y. [i1 down]…[i1 down, up, down] Yeah, anywhere around about there is going to be fine.* (p18)

> *So the middle one [i3MF] is usually in the middle for me anyway.* (p10)

Described perceptual changes as well as evaluations during anticipatory adjustments referred to issues described prior to the adjustment.

> *Oh, it’s very shrill. So, I need to kill that a bit. (…)*

> *[i2HF down] I need to move that down.*

> *[i2LF up] I think that’s about right…--no, a bit shrill.* (p17)

> *So, that’s interesting, each time they start off--it starts off pretty shrill. [i3LF up] so I always give it a bit of bass.* (p08)

When issues could not be resolved, verbalisations reflected a process of re-evaluating or reweighting of issues.

> *Och…it’s that background noise -- I can’t stand it.*

> *[i1 down] Oh no, and that gets too loud.*

> *[i1 up] You see, when you adjust that, you’re actually…(waves hand) not changing the background noise.*

> *[i1 down] Yeah, that’s probably a bit better. But it’s a wee bit loud…As soon as you make it--As soon as you try and make it not as loud for his voice, you’re still getting that noise in the background.*

> *(…) I think you have to just try and ignore the background noise and concentrate on his voice.*

> *[i1 up]…[i1 down, up] Yeah, I’m going to go back to there.* (p12)

Similarly, the search for solutions was often accompanied by crossing over to a more exploratory mindset.

> *I have a go, going up the way. [i3HF up]*

> *Actually that’s better! [i3HF up] I hope I’m not messing up your results. [i3HF down, up] [pauses playback] See, when he says ‘I can’t bear disgrace’, in all the other trials he’s been annoying me as he’s not been saying ‘disgrace’ clearly. I heard that as clear as anything.* (p25)

> *[i3MF down] I wonder if the mid frequencies will--…*

> *[i3MF down] Oh, that’s interesting, reducing that seems to have reduced the background noise quite a bit, I can hear him more clearly.* (p09)

#### After the adjustment

The exploratory approach involved evaluating the adjustment endpoint hedonically against prior settings. Self-adjustment endpoints were considered the best possible setting from the options available, and evaluations were moderated by the exploration (i.e., expectations tempered by immediate prior experience).

> *So, yeah, I’m quite happy with that, I could quite easily listen to that.* (p08)

> *[i1 up] But all that background noise is awful, [i1 down]…awful, awful. [i1 down] And there’s absolutely no way to get that one to feel comfortable at all, none whatsoever.* (p14)

> *[i3HF down a notch] That’s probably as good as it’s going to get — Cause it got all that background stuff.* (p05)

The anticipatory approach involved evaluating whether adjustment resolved the identified issues in the initial sound or during adjustment. Positive and negative evaluations also contributed to building experience with the adjustment task, with insights gained from successful or unsuccessful outcomes subsequently applied in later trials.

> *I got the bass. Got the clarity of voice. I think that’s ([incomprehensible:] it).* (p23)

> *That gives it a richness (…)richness and a bit of depth…without being too bassy.* (p13)

> *I was surprised that reducing the middle one, by reducing the middle one, it actually made it sound clearer.* (p09)

### Theme 2: Perceptual changes and reactions when adjusting sliders

During adjustment, verbal reports described interactions, resulting perceptual changes and evaluations of these changes, potentially leading to further interaction (centre of Figure 3). For example, the sequence

> *[i3LF up] and as I push it up, the volume goes up. And that’s a bit much for me.* (p10)

**Figure 3.**
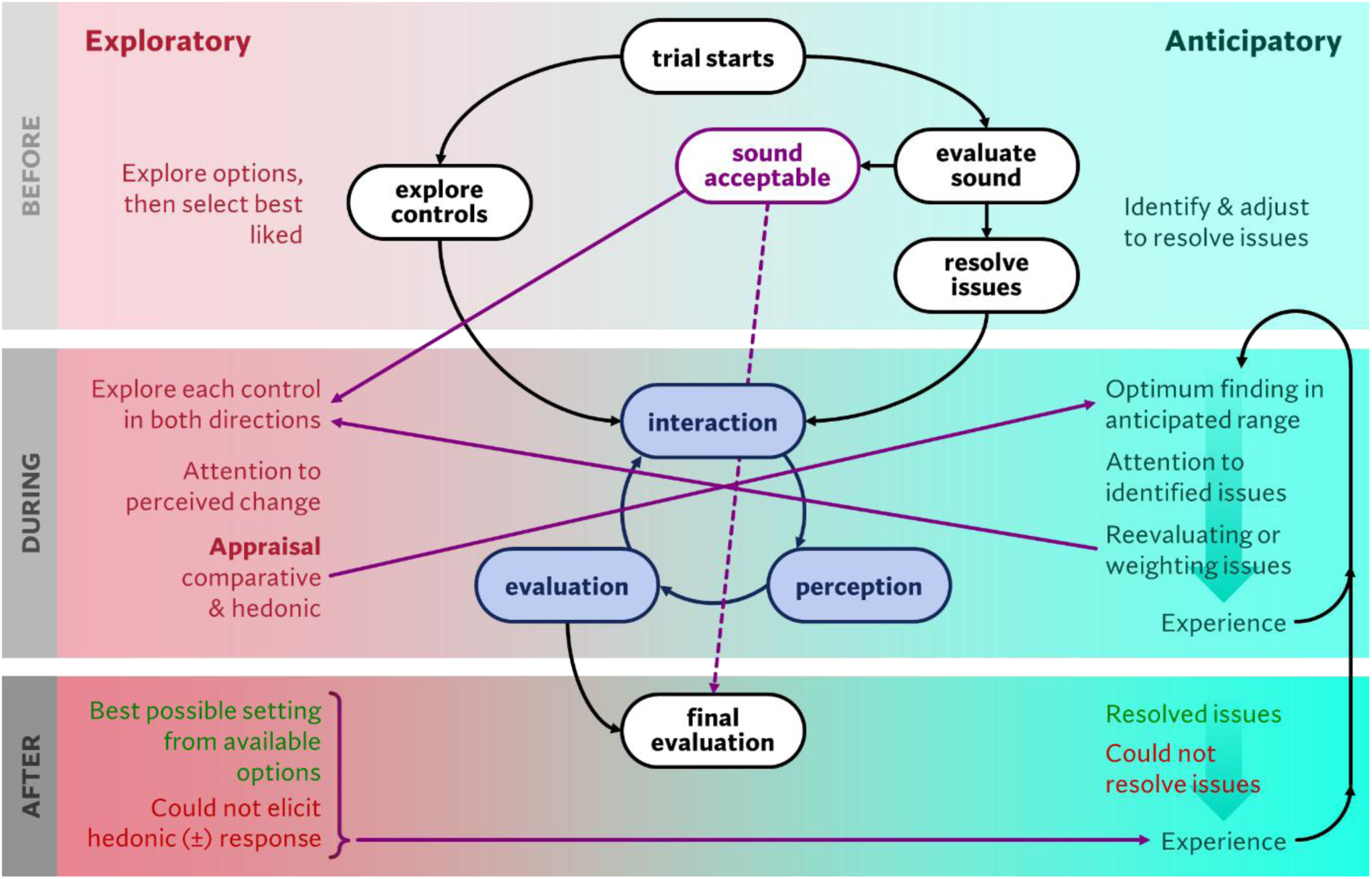
Suggested model illustrating a spectrum of anticipatory (left) and exploratory (right) FGR self-adjustment archetypes, including typical crossover (purple lines) experience (black lines) pathways across three stages of self-adjustment (before - top, during - middle, and after 一 bottom).

> *describes the interaction (as I push it up), the perceived change (the volume goes up) and an evaluation (that’s a bit much for me). However, reports were not strictly adhering to this sequence, possibly due to the speed of adjustment and thoughts compared to verbalising them. For example,*

> *[i2LF down] Oh, no, no, no, no. Too much background.* (p26)

Here, the negative evaluation, or rather how the perceptual change is perceived (*no, no, no, no*) is verbalised prior to describing what perceptual change was (*Too much background*). It can be however assumed that both a percept and an evaluation occurred, but both may not be distinct, in particular if the sound was markedly unacceptable.

Further analysis and identification of sub-themes of Theme 2 was focussed on verbalisation of perception and evaluation of adjustments, rather than the time-order and process of their occurrence. Within Theme 1, participants’ descriptions of slider functions were organised by interface and direction of slider movement from the previous point, irrespective of magnitude. Verbalisations were not organised by stimulus type, although certain interpretations were specific to the content of the stimulus. This decision was based on the observation that, at the group level, there were no (or negligible) systematic differences in adjustments as a function of stimulus type. Additionally, once a slider was interpreted as controlling a particular dimension (e.g., *bass*), that interpretation tended to remain consistent across subsequent trials. Nevertheless, stimulus types did surface in participants’ descriptions when relevant (e.g., references to ‘less background noise’ appeared only in response to speech-in-noise stimuli). Such cases were therefore categorised under *content* and *descriptors × content*. The following subthemes were identified with examples for each:

- **Slider functions**: what a slider does (e.g., *controls bass* or *treble*)

> *[i1 up, down, up] That’s just an overall balance between bass and treble.* (p15)

> *[i2HF up] That’s treble boost, I would say. For intelligibility I want to put that up a wee bit.* (p15)

> *[i2HF up] — see what this one does….[i2HF up, down] Ahh, that’s bass and treble.* (p06)

- **Sound descriptors**: adjectives used to describe the sound (e.g., *tinny* or *clear*)

> *[i1S up] it’s clearer, but it’s still too loud.* (p14)

> *[i3MF up] It’s a bit tinny.* (p16)

> *[i3HF down] That’s richer…richer.*

> *[i3LF down] Very clear tone.* (p04)

- **Content**: how an adjustment changes what can be heard (e.g., *more background noise*)

> *[i1 up] And the background is coming through a wee bit more there.* (p13)

> *[i3HF down] This is--this is losing some of the music* (p27)

- **Descriptors × content**: sound descriptors that also include content (e.g., *warmer voice*)

> *(…) Just enhancing his voice [i2LF up] Yeah. Making it warm.* (p18)

> *[i3LF down] I’m finding this voice very sharp.* (p27)

- **Hedonic responses**: characterised by a change of how pleasurable the sound is (e.g., *unpleasant* or *much better*)

> *[i3HF up] I like that, that’s nice.* (p19)

> *[i3LF down] That’s more pleasant down there.* (p11)

> *[i1 down up] Oh, increasing it makes it really unpleasant.* (p09)

#### Agreement between adjustments and thoughts

There was some agreement between participants’ actions and verbalisations while adjusting. Boosts in any part of the spectrum were described as *louder* or *more [descriptor/content]*; attenuations were described as *quieter*, including potential synonyms (e.g. *softer*) or *less [descriptor/content]*. Spectral tilts in adjustment were identified as *more/less bass* (and synonyms, e.g. ‘deeper’), and/or *less/more treble* (and synonyms, e.g. ‘more high-pitched/shrill’). The description of relative gain, in particular low to high frequency, was observed both in i1 and with attenuation in i2 and i3. That is, attenuations lead to a description of the unchanged part of the spectrum (e.g., a reduction in treble was described as *more bass*). Descriptions of spectral tilts occurred sometimes in tandem with descriptions of perceived loudness.

Agreement was also found when hedonic or affective responses were linked to particular content and adjustment direction. For example, for speech in noise, a decrease in frequency-gain was described as *less* (i.e., a reduction in background noise) and elicited a positive hedonic response, and an increase, described as *more*, elicited a negative response. In the examples below, of note is the discovery of context-dependent functionality for i3MF, resulting in a positive affective response.

> *[i2LF down] Yeah, that’s better. Yeah, the background noise is gone a bit.* (p12)

> *[i1 down] That brings up the sound in the background, which isn’t good.* (p01)

> *[i3MF down]…Hmmm…That’s actually cutting out a lot of the background noise.*

> *[i3MF down] That’s--I prefer that actually.*

> *[i3MF up]…The noise is increasing but not the clarity. [i3MF up] Ow!*

> *[i3MF down] Hmm…quite like that. It has cut down the background noise.* (p25)

> *(…) [i2LF up] This is better cause it’s bringing the music forward.*

> *[i2HF down] This is--this is losing some of the music (…)* (p27)

#### Challenges describing sound

Slider function and sound descriptors often overlapped, with participants using the same words for both (e.g., *bass* t *bassy, more bass*). Not all participants used sound descriptors readily except for “loudness.” Some verbal reports suggested that accessing appropriate vocabulary was challenging. This was indicated by, for example, reliance on hand gestures to convey meaning, explicit word searching or exasperation:

> *… and letting the percussion (hand gesture, moving both hands outward in opposite directions in front of him ∼’large’/’expansion’)…to give it…the hight, yeah, that’s what it sounds a bit.* (p18)

> *If I do that actually, if I do that up there (points at i3LF), it’s loud, but if I take it further down, it’s a wee bit distorted, for the lack of a better word.* (p24)

> *[i3LF down, up] Bass! That’s the word I was looking for. (p03, last trial)*

In particular, describing the function of i3HF sometimes posed challenges, additionally indicated by, for example, reflexive questioning, hesitation and surrender:

> *[i3HF up, down] -- No? Up and down.*

> *[i3HF up, down, up] And…[i3HF down] it does?…There’s more bass down there, but that’s not [incomprehensible] it. No, I don’t know. I don’t know what that one’s doing.*

> *[i3HF up, down] But that’s easier listening.* (p06)

There were indications that some of these challenges could have been due to differences being perceived as small and hence not meaningful in certain participants:

> *(…) it’s ok, but if i take it a little bit further, it’s not a lot of difference between that one [points at i3HF centre of slider] and that one [points further down]. It’s not a lot of difference, really. So I put it back up.* (p24)

### Observations of adjustment behaviours

#### Bracketing

A bracketing strategy is deliberately (initially) over- and undershooting to find the appropriate setting (e.g., self-adjustment endpoint) between these extremes (cf. Stevens, 1959). Bracketing has been observed in past studies (e.g., Goβwein et al., 2024); however, verbal reports of using this strategy were rare. Instead, participants’ verbalizations typically described perceptual impressions of ‘too much’ or ‘too little’, without indicating a prior intention to explore an acceptable range through bracketing.

*[i2HF down]…[i2HF up] That’s clearer, his voice is clearer there, but it’s louder as well. [i2HF down] Now, when I take it down there, it gets fuzzy. So I would leave it in the middle. [i2HF up]* (p07)

#### Trade-offs and reactions to adjustment interfaces

Situations requiring participants to negotiate a trade-off between different optima, either by prioritising certain adjustment criteria or settling on a compromise, were predominantly observed in connection with the speech-in-noise stimulus.

> *[i2LF up] As you go up it’s just--(shakes head)…[i2LF down, up] is almost as if you can’t win.*

> *Cause if you go up, you hear all that background stuff…*

> *[i2LF down] If you go down, it’s not as clear.*

> *[i2HF up, down] (mumbles:) Don’t like that either.*

> *[i2LF up] That’s probably as good as it’s going to get.* (p05)

> *[i1 up] You see, when you adjust that, you’re actually (waves hand) not changing the background noise.*

> *[i1 down] Yeah, that’s probably a bit better. But it’s a wee bit loud. As soon as you make it--As soon as you try and make it not as loud for his voice, you’re still getting that noise in the background.*

> *[i1 up] That’s probably better.…I think you have to just try and ignore the background noise and concentrate on his voice. [i1 down, up] (…) Yea, I’m going to go back to there.* (p12)

#### Issues with task

Some participants expressed that the task felt repetitive. This included experiences from previous tasks regarding issues with the initial sound as well as how to adjust to improve them.

> *Once more: too loud for me.* (p14)

> *Just the same; I’m looking for a basser--[i2LF up] That’s too loud and that’s too bassy.* (p20)

> *Again, his voice is too thin.* (p22)

One participant raised concerns that she could not focus on both tasks, adjustment and thinking aloud, simultaneously. This indicated, at least for this participant, that concurrent TAP did consciously interfere with adjustments.

> *[HF up, down] You would never think that trying to do the two things…would affect it, you know, like, to work as one. You tend to concentrate on one thing only, to move this, to make it right, and then you realise, you’re not doing that, you know.* (p02)

#### Intelligibility of speech in stimuli

Participants noted that the speech stimuli were easy to understand attributing this to the speaker’s way of speaking.

> *I like this man’s voice. This man’s voice is easy…is easy to understand and to hear.* (p04)

> *[i3LF down] See this guy, you can listen to this guy, y’ know. Once you got it sorted, y’ know. It’s--aye, good speaker.* (p21)

#### Difficulty deciding on slider endpoints

Some participants expressed difficulty deciding on a slider endpoint. When expressed without additional context, these verbalisations are difficult to interpret, for example:

> *It’s quite difficult to decide where the best situation would be.* (p01)

Another participant expressed that adjustments of the FGR lacked relevance and did not lead to increased preference.

> *You can make the sound different…But that doesn’t--doesn’t mean…it’s--errr…you prefer it more. You can just make it different -- doesn’t make much difference. I suppose it depends on what the sound is eventually coming to your brain as. It’s with the whole picture, that doesn’t matter an awful lot in that process.* (p07)

This participant questions whether adjustments lead to meaningful improvements, implying that other factors than FGR may affect auditory experience; the perception of acoustic change may be less important than other cognitive or contextual factors influencing how the brain interprets sound. Without being stated explicitly, possible factors could be attention, listening context, or emotional state. This ‘different not making a difference’ perspective, while singular, challenges the underlying assumption of self-adjustment that participants can link acoustic changes to perceptual preferences. That is, that even perceivable changes may not be meaningful to the user (cf. McShefferty et al. 2016). Instead, it points to a more complex interaction between sound, cognition, and context, where the efficacy of self-adjustment is not solely determined by the technical outcome of the adjustment, but by how that change is perceived and evaluated within a broader psychological and situational framework. A counterexample expresses how this level of engagement promotes careful listening, whereas adverse sounds (too loud, too noisy) has the opposite effect:

> *So, you got to adjust it to get involved, to listen. Because you don’t want to listen when it is too loud or when there’s background.* (p21)

These types of thoughts, however, can be largely considered high-level verbalisations (i.e., providing reasoning) which have been criticised as including non-task information concurrently attended to by the participant (Fox et al., 2011). This may introduce reactivity, shifting participants’ focus from searching the most perceptually preferred option to choosing an option that is best supported by their articulated reasoning (Wilson & Schooler, 1991).

### Self-adjustment with and without TAP

All participants previously completed the same self-adjustment task 4-22 weeks earlier without TAP (Benecke & Whitmer, 2026). Compared to their previous session, the median trial duration of the self-adjustment with TAP was longer by a factor of 1.5-1.9 (26.5, 36.9 & 44.4 s for i1, i2 & i3, respectively, compared to 16.1, 19.5 & 24.4 s in the previous session), which was expected as the concurrent verbalisations generally slow any process (Ericsson & Simon, 1993). The self-adjustments endpoints produced during TAP could potentially differ from the previous self-adjustment endpoints due to difficulty focussing on the adjustment while thinking aloud (i.e., divided attention), reactivity to verbalised thoughts [e.g., stating a desire for more bass, then exaggerating the bass beyond previous preference], accommodation to the sound of their own unamplified voice and the time delay between the first and second session. Participants’ mean adjustment endpoints per slider in the previous session are compared with those in the current session with TAP in Figure 4. At the group level, adjustments were relatively consistent across sliders i1, i2HF, i3MF, and i3HF with median differences < 1 dB. However, the gains significantly increased with TAP for i2LF and i3LF slider [median Δ = 3.2 & 3.7 dB, respectively; *z*(36) = 2.92 & 4.13, respectively; both *p* < 0.05]. These differences agree with the aforementioned reactivity [e.g., *I like the bass* (p03, p20, p22)]. At the individual level, most individual adjustments were within the standard deviation of their previous session (dashed red lines in Figure 4). In the current study, the focus was not the ends 一 to where the participants adjusted 一 but rather the means 一 what they thought about during the process.

**Figure 4.**
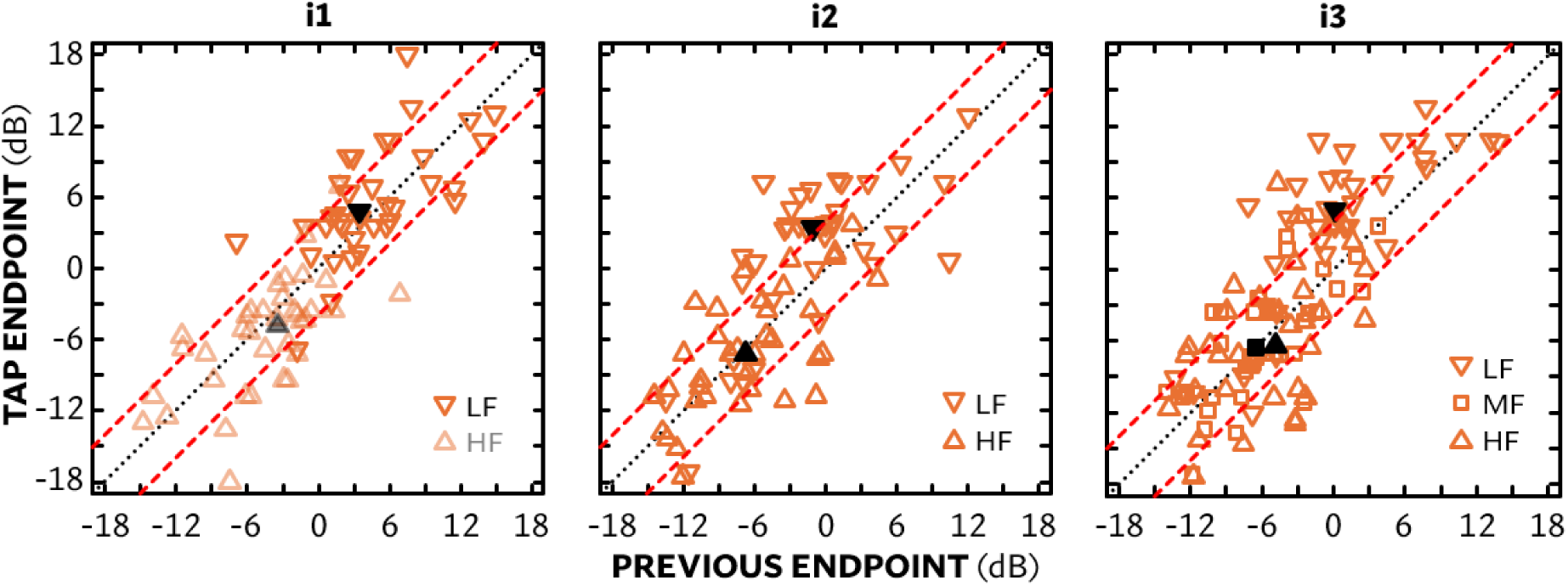
Individual mean self-adjustment endpoints for TAP (current) session for each interface (panel) as a function of previous session mean endpoints (Benecke & Whitmer, 2026). Shapes refer to different sliders: downward triangles = low-frequency (LF); squares = mid-frequency (MF); upward triangles = high-frequency (HF). Filled black symbols refer to medians. Dashed red lines indicate ±2.8 dB (std. deviation in previous study) from zero difference in endpoints (diagonal dotted line). For i1 (left panel), the HF endpoints are the inverse of the LF endpoints.

## DISCUSSION

This study is, to our knowledge, the first using concurrent TAP to gain insights into the approaches guiding FGR self-adjustment. Previous studies of self-adjustment that have analysed behaviour (e.g., Gäβwein et al., 2024)have relied on making speculations on the aims and motivations for adjustment using solely quantitative data. The analysis revealed that FGR differences in self-adjustment can be, with the possible exception of i3HF differences, meaningfully described, and that the process is navigated along a spectrum of exploratory and anticipatory mindsets. Participants’ verbal reports contain mixed forms or crossovers from one archetypical mindset to the other. The archetypes are useful in understanding the different mindsets used, possibly by the same individual during the same self-adjustment session, when navigating the adjustment process.

At one end of the spectrum, the exploratory archetype reflected an approach motivated by interaction with the adjustment interface itself. Exploratory verbalisations described slider functions, and FGR differences were evaluated hedonically against immediately prior percepts. Exploratory self-adjustment can be characterised as finding an optimum within available options. At the other end of the spectrum, the anticipatory archetype was characterised by verbalisations evaluating the initial sound and explicit goal setting for the adjustment. Drawing on participants’ prior experience -- from immediately preceding TAP trials to their previous session without TAP providing ample exposure to the same interfaces to potentially their own long-term experiences -- the anticipatory archetype engaged in optimum-finding within an expected FGR range. Evaluation of changes and decisions to stop adjusting were guided by the initial evaluation and the predefined adjustment goal.

When participants were expressing perceptual changes or affective reactions while adjusting a slider, these expressions generally agreed with the spectrum and direction of gain change. A few participants expressed that they were unsure about the function of i3HF, or what differentiated i3MF from i3HF. Not all participants gave detailed reports on perceptual changes, and some expressed how it was difficult to find the right words. At a group level, the self-adjustments made while thinking aloud agreed with these participants’ self-adjustments in a previous session save for 3-4 dB increases in independently controlled low-frequency gain in i2 and i3.

### Contrast to descriptor mapping in paired comparisons

Sound descriptors, when used in the current TAP, were largely in agreement with findings described by ‘expert systems,’ the sound descriptors reported by audiologists to be used in patient complaints leading to hearing aid personalisation (Jenstad et al., 2003). That is, the TAP participants used the same terminology as suggested by clinicians when describing the sound before or after a change in FGR. This contrasts with the lack of reliable and consistent descriptor-to-FGR mapping in previous results evaluated quantitatively in a pairwise comparisons study (Caswell-Midwinter & Whitmer, 2021). A range of methodological differences may account for this disagreement in findings. The previous study quantitatively measured intra- and inter-rater reliability for the descriptors, whereas the descriptors in the current TAP were evaluated qualitatively; it did not matter how many participants used the descriptors, only that they were used. The previous study used repeated unforced-choice pairwise comparisons of fixed FGR changes up to ±12 dB to elicit choices from a fixed number of descriptors from Jenstad et al. (2003), whereas the current TAP study used the method of adjustment offering a greater range (±18 dB), and sound descriptors were freely verbalised. By using the method of adjustment, participants had active control over the adjustment process, including the amount of change, duration of exposure, and the timing and sequence of adjustments. These TAP results during adjustments suggest that active control of the sound by the participant, as opposed to passive pairwise comparison, can promote the operationalisation of sound descriptors. Additionally, this active control and how it leads to anticipatory behaviours during the process poses a counterargument to the critique of self-adjustment by Jensen et al. (2019). While there is an inherent this-or-that simplicity in a pairwise decision, the trial-and-error in self-adjustment can promote greater understanding of what can be achieved in personalisation.

### Exploratory-anticipatory model in relation to quantitative behaviours

The identified exploratory and anticipatory archetypes for navigating self-adjustment help contextualise the participant interactions observed in Goβwein et al.(2024). That study grouped participants depending on how much they explored the available parameter space and discerned clusters of behaviour such as ‘full-on browsing’ (i.e., exploring the full range available) or ‘carefully browsing’ (exploring a narrower range). This wide vs. narrow range difference fits into the current exploratory-anticipatory model. An exploratory approach can result in a wide adjustment range, testing the interface’s capacity or employing a bracketing strategy, whereas an anticipatory approach can result in a narrower range with adjustments guided by the intention to change the sound. For example, if a participant anticipates a particular axis of the interface to yield quieter settings, they may intentionally search in that region and avoid regions they expect to be associated with increased loudness. However, when analysing and interpreting interaction patterns, it is important to consider not only the adjustment style (or archetype), but also the starting point, the adjustable range, and the perceptual salience of frequency-gain changes within that range.

### Integrating TAP into clinical practice

Participants’ responses generally reflected a positive attitude towards self-adjustment, as has been reported in numerous previous studies (e.g., Elberling & Vejlby Hansen, 1999; Dreschler et al., 2008; Nelson et al., 2018; Boothroyd et al., 2022). Boothroyd et al. (2022) demonstrated that the majority of participants preferred shared decision-making between audiologist and themselves when fitting hearing aids. A substantial proportion of responses indicated a preference for either ‘Me with input from a professional’ or ‘A professional with input from me’ (ibid., Fig. 8). The nature of the input provided by the patient is not clearly defined, but the content and quality of this input could significantly influence the effectiveness of shared decision-making. If patients use a self-adjustment tool for frequency-gain personalisation in the clinic, a concurrent TAP approach would help assure that the content and quality of the shared decision-making is better associated with the personalisation by directly coupling patient reaction to a sound change to FGR and/or other parameter changes. Even if the concept may not lead to changes in FGR settings compared to an initial audiometric response, it may be a way for the hearing aid user to explore and therefore understand the options available to them and, crucially, promote greater ownership in the process.

This approach also has the potential for audiologists gain a better understanding of the patients’ unique vocabulary used, as well as the limits of acceptability for certain settings, and with what aspects of self-adjustment they may struggle. The exploratory-anticipatory model can be used as a guide for both patient and clinician. Future research should focus on developing the exploratory-anticipatory model as a tool for patient and clinician, providing a structured framework in which such sessions could take place. Another aspect warranting further study is the selection of appropriate stimulus materials that are available but relevant to the patient. Recorded media, such as music, podcasts or film/TV as stimulus could offer the advantage of allowing patients to select stimulus material themselves but may lack realism and therefore generalisability to everyday listening situations. In particular, voice recordings that typically feature well-articulated speakers or minimise room acoustics differ in pace and content from natural, everyday speech.

### Implications for self-fitting

Self-adjustment archetypes have important implications for devices which users set up independently of a clinic, that is, self-fit. Effective guidance on self-fitting should account for different adjustment styles. Having a self-fitting tool that sets adjustment goals may not work with someone in an exploratory mindset; however, they may benefit from a more structured approach including bracketing. In the same vein, for an individual in an anticipatory mindset who evaluate against initially determined issues, a tool encouraging them to explore may help them to discover different settings (e.g., distinguish between issues attributed to the stimulus/sound or resulting from amplification). To evaluate adjustments, it may be helpful to provide access to initial settings, for example include a before and after comparison in self-adjustments could be useful. An issue for future research to resolve is determining efficiently and robustly when someone is in an exploratory or anticipatory mindset based solely on their behaviours.

### Limitations

The thematic analysis was inductive and reflexive, and as such departing from positivist rigour. As a result, interpretation of the qualitative data was subjective, no inter-coder agreement measures were applied. Likewise, no quantitative methods such as assessment of code frequency were applied to the data. These are not limitations, but rather design decisions based on the notion that individual verbal reports may not have been complete, due to participants interacting faster than the time required to verbalise thoughts and the varying degrees to which they were comfortable with and amenable to thinking aloud. One known limitation in the analysis was that no external validation, such as participants reviewing their own transcripts, was conducted. This step was omitted to avoid overburdening participants who had already committed to two intensive sessions.

The task, although reduced to only nine adjustments being unique combinations of stimulus and interface, was still perceived as repetitive. While some repetition may be necessary to afford chances to talk about otherwise missed impressions, starting self-adjustments from different baselines or to more varied or novel stimulus types could have prevented perceived repetitiveness, but also possibly broadened results or posed difficulties with identifying themes. It is also possible that the prior exposure to the self-adjustment procedure and interfaces heightened the sense of repetition verbalised by some. With that prior exposure, it is uncertain whether findings generalise to users who are new to self-adjustment and therefore lack prior experiences that might help them build anticipation.

While none of the participants opted out, and none of the participants fell completely silent throughout the trials, there were instances of trials without verbalisation, often due to the participant moving on to the next trial just before being reminded to keep talking. Thinking aloud can be challenging for some individuals, in particular if trying to only obtain low-level verbalisations (i.e., inner thoughts held in short-term memory present in word form or translated to word form; Ericsson and Simon, 1984). Higher-level verbalisations that are not currently in attention and involve information stored in long-term memory may not accurately reflect in-the-moment thought processes, and participants may be constructing plausible reasoning retrospectively (ibid.). These retrospective thoughts may introduce reactivity, affecting thoughts as well as behaviours, therefore affecting the validity of the resulting model. The relative increase in low-frequency gain in the current session relative to the same participants’ past session (Figure 4) supports the possibility that adjustments were reactive to participants’ expressions. However, none of the features of the exploratory-anticipatory model described above would be affected by this, only the endpoints. Further study is needed to see if potentially reactive endpoints are more or less preferred than those without TAP.

## CONCLUSIONS

Using TAP, a common HCI method, to explore the self-adjustment of hearing-aid gain has provided valuable insights into individuals’ perceptions of gain change and approaches to personalisation. These perceptions and approaches form a qualitative model of self-adjustment that characterises individuals’ behaviours along a spectrum from anticipatory to exploratory strategies throughout the adjustment process. Movement across this spectrum may be influenced by prior exposure to and experiences with the self-adjustment interfaces and stimuli used in the experiment as well as individuals’ aims in interacting with the self-adjustment tools provided. Additionally, we found that TAP is feasible during self-adjustment, though it is constrained by limitations such as the fast-paced nature of the task and general challenges associated with TAP (e.g., concerns about validity, reactivity, and inclusivity). Since the verbalisations were coherent with participants’ interactions, the validity of the results can be reasonably assumed. With some adaptation, TAP has the potential to be a useful and effective tool in connection with self-adjustment in the clinic where it could serve as a basis of a conversation and shared decision-making between audiologist and patient. This may also shift the focus away from speech benefit and troubleshooting to empowering hearing-aid users to approaching ‘better’ without the need to explain, but a tool to demonstrate what better means. Given the success here using TAP in the laboratory, future studies should apply TAP with the self-adjustment of hearing aids using hearing-aid apps in realistic, personally relevant environments to further develop this model for the clinical, remote and self-guided personalisation of hearing-aid gain. Future research should also use TAP to investigate the thought processes on numerous initial settings.

## Data Availability

Anonymised study data is currently being prepared for repository submission

## ACKNOWLEDGMENTS

This work was supported by funding from the Medical Research Council [grant number MR/X003620/1]; and GN Hearing A/S. Thanks to Prof. Graham Naylor, Gregory Olsen and Dr. Qi Yang for helpful comments throughout this work, and to Andrew Lavens and David McShefferty for technical assistance.

## SUPPLEMENTAL MATERIALS 1. VERBOSE MODEL SCHEMATIC

Figure S1 shows an alternate view of the model with data (i.e., example quotes) in place of subthemes at each stage of the self-adjustment process. Note that many of these quotes were accompanied by gestures that provide additional context but were omitted due to space.

**Figure A.1.**
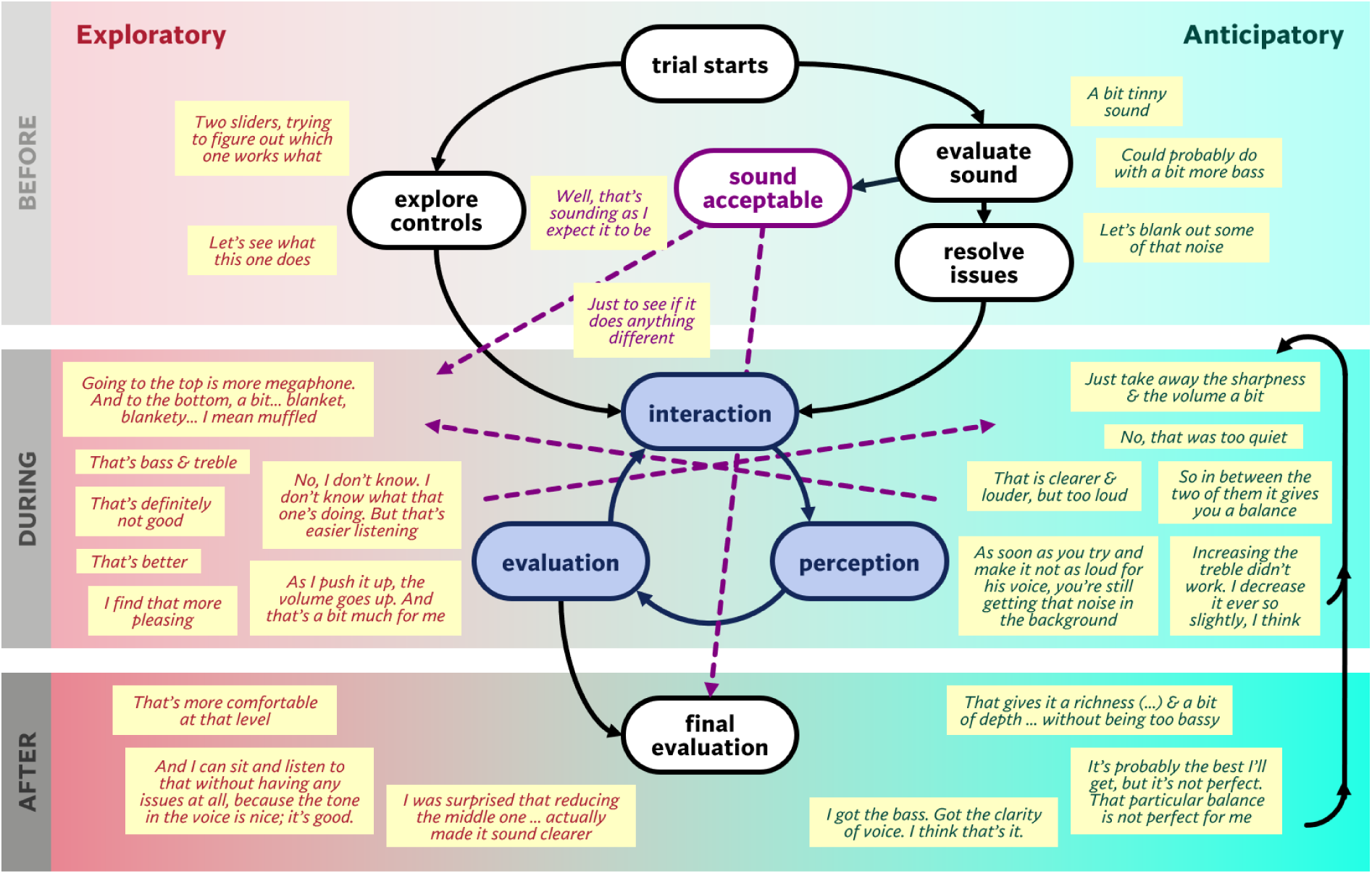
Qualitative model of self-adjustment using exemplar quotes to demonstrate the spectrum of anticipatory (left) and exploratory (right) FGR sef-adjustment archetypes across three stages of self-adjustment (befOre - top, during 一 middle, and after 一 bottom).

## SUPPLEMENTAL MATERIALS 2. TAP TRANSCRIPTION STYLE GUIDE

When transcribing and annotating utterances and actions during TAP, we’re aiming for verbatim transcription of what participants expressed verbally and non-verbally, such as hand gestures and other body language. The goal is to have a transcript that can replace the video, that is it captures all essentials to interpret when coding.

### Transcribing verbal content

Transcripts do not need to follow Jefferson transcription but should capture and convey the meaning or context; that is, how something is said. The following will serve as style guide on how to deal with particularities of spoken language (fill words, pauses, interrupted speech, mumbling):

- Whenever fillers, exclamations or giggles can be heard, write them down phonetically (*err, hm, um, mmh, uh, ah, oh, hahaha* etc.)
- For pauses in speech use ‘…’ (aka ellipses). Rather than defining a pause by duration in seconds (these vary and may depend on a participant’s speech rate) use your own judgement when experiencing an unusually long silent gaps in speech; if not sure about a pause, mark in brackets ‘(…)’
- *example: “I think—…ummh…Bass! That’s the word I was looking for.”*
- Interrupted speech/sharp stop or stuttering 一 ‘一’
- *example: “I—…what does— [i3HF up, down] I’m not quite sure what it does.”*
- Full stops or other punctuation to end a sentence 一 use you best judgement to find out emphasis (‘!’) or raised voice by the end to make a statement rather sound like a question (‘?’). If phrasing is suggesting question, but voice is not raised at the end, feel free to use (.) or other suitable punctuation. (“*Right? Right*.”)
- Mumbling or otherwise incomprehensible f use brackets () and give reason why it was incomprehensible and then best guess denoted with a ‘?’. If it’s impossible to guess denote ‘(incomprehensible)’.
- *example: “(mumbles (incomprehensible)) back (mumbles: and?) hearing more about Sherlock.”*
- Tone of voice, if relevant and text understanding would be difficult otherwise. *example: “(jokingly:) Oh no! I think I don’t want to listen to that story ever again.”*
- If important words are missing for context, feel free to annotate using ‘[word that is missing implied from context, e.g., what happened prior]’. This may help identifying the context when the text appears out of context, such as a coded snippet.
- *example: “Right, try another one [trial].”*
- In the same vein as above, you might encounter situations in which a participant refers to a previous interaction or trial. If so, feel free to give context by explanatory text in brackets 一 ideally, we may get together to get consensus on these hopefully few cases.
- Interruptions in playback 一 annotate in square bracket ‘[pause/resume playback]’
- Interruption from experimenter: include in transcript if it is short and/or participant refers to it, in square brackets with ‘[E: verbatim transcript of what experimenter said]’. If there is a longer passage in which talking-aloud is explained to the participant, it is ok to omit annotation ‘[E: (explains experiment task)]’.
- If the participant’s words don’t make sense, or the grammar is wrong, do not correct 一 if it’s obviously wrong, but this is what they said add ‘(sic)’, even though the s stands for *scriptum*.

### Annotating gestures

#### a) Interactions with sliders

Note when participant interacts, that is in most cases, moves a slider, and the direction(s) the slider is moved in (e.g., up, down, or in case of a series of smaller up and downs: wavering). Annotate that interaction in square brackets within the transcript of utterances, annotated interaction does also signify speech pause.

Sliders of each interface are named as follows:

- interface 1, single slider > i1S;
- interface 2, LF (left/low frequency) and HF (right/high frequency) T i2LF, i2HF;
- interface 3, LF, MF, HF (left to right, low, mid, high frequency)玲 i3LF, i3MF, i3HF *example: “(mumbles: What does this?) [i3MF up, down] Oh, that’s much clearer.”*

#### b) Body language/hand gestures

If hand gestures, facial expressions (if visible) or general body movements are used in a meaningful way, e.g., gesture replacing words. There is no need to annotate all gestures, but only those that convey meaning, and make utterances less ambiguous or clearer.

- Describe hand gesture in text in square brackets at moment of occurrence *example: “I find this background quite distracting [talking hand gesture]”*

As framework, a classification of hand gestures as proposed by McNeill (1992) may be helpful:

- beat: reflect or emphasise aspects of speech such as tempo
- deictic: pointing gestures, used to refer to at real item, indicating directions or referring to something in the (immediate) past
- iconic: representing something physical, such as a person or an object
- metaphoric: gestures representing an abstract idea

Beat gestures do not need to be transcribed but may help in deciding on punctuation (e.g., when to use ‘!’). Deictic annotation is useful only when participants are referring to sliders in an ambiguous way (e.g., *“not sure about that one [pointing at i3HF]”*); please annotate which slider they are referring to only if it is a slider they have not interacted with in the same utterance. Similarly, include icons and metaphoric gestures only if they add to what was said, but not if participant is expressing the same verbally and with gestures (e.g., if describing the sound as ‘rounded’, do not annotate if they make a ‘globe or ball shape’ iconic hand gesture).

## REFERENCES

Benecke, J., & Whitmer, W. M. (2026). Does the method matter? Evaluating the effectiveness, efficiency and ease of hearing-aid gain self-adjustment [pre-print]. MEDRXIV/2026/355463.

Bentler, R. A., & Cooley, L. J. (2001). An examination of several characteristics that affect the prediction of OSPL90 in hearing aids. Ear and Hearing, 22(1), 58-64. l0.1097/00003446-200102000-00006.

Bjerg, A. P. & Larsen, J. N. (2006). Recording of Natural Sounds for Hearing Aid Measurements and Fitting, 0rsted, Denmark: Danish Technical University (DTU), Acoustic Technology.

Boothroyd, A., & Mackersie, C. L. (2017). A ‘‘Goldilocks’’ Approach to Hearing-Aid Self-Fitting: User Interactions. American Journal of Audiology, 26(3 Suppl): 430–435. 10.1044/2017_AJA-16-0125.

Boothroyd, A., Retana, J., & Mackersie, C. L. (2022). Amplification Self-Adjustment: Controls and Repeatability. Ear and Hearing, 43(3): 808–821. 10.1097/AUD.0000000000001141.

Braun, V., & Clarke, V. (2006). Using thematic analysis in psychology. Qualitative Research in Psychology 3: 77–101.

Braun, V., & Clarke, V. (2021). Can I use TA? Should I use TA? Should I not use TA? Comparing reflexive thematic analysis and other pattern-based qualitative analytic approaches. Counselling and Psychotherapy Research, 21(1): 37–47. 10.1002/capr.12360.

Braun, V., & Clarke, V. (2022). Conceptual and design thinking for thematic analysis. Qualitative Psychology, 9(1): 3–26. 10.1037/qup0000196.

Byrne, D., & Dillon, H. (1986). The National Acoustics Laboratories’ (NAL) new procedure for selecting gain and frequency response of a hearing aid. Ear and Hearing 7(4): 257–265.

Caswell-Midwinter, B., & Whitmer, W. M. (2019). Discrimination of Gain Increments in Speech. Trends in Hearing 23: 2331216519886684. 10.1177/2331216519886684.

Caswell-Midwinter, B., & Whitmer, W. M. (2021). The perceptual limitations of troubleshooting hearing-aids based on patients’ descriptions. International Journal of Audiology, 60(6): 427–437. 10.1080/14992027.2020.1839679.

Cunningham, D. R., Williams, K. J. & Goldsmith, L. J.(2001). Effects of providing and withholding postfitting finetuning adjustments on outcome measures in novice hearing aid users: A pilot study. American Journal of Audiology 10 (1): 13–23. doi:10.1044/1059-0889(2001/001).

Debevc, M., Žmavc, M., Boretzki, M., Schüpbach-Wolf, M., Roeck, H.-U., Khan, A., Koubatis, A., Jezernik, S., & Kožuh, I.(2021). Effectiveness of a Self-Fitting Tool for User-Driven Fitting of Hearing Aids. Int. J. Environ. Res. Public Health, 2021, 18, 10596. 10.3390/ijerph18201059.

Dreschler, W. A., Keidser, G., Convery, E., & Dillon, H. (2008). Client-based adjustments of hearing aid gain: The effect of different control configurations. Ear and Hearing 29(2): 214–227. 10.1097/AUD.0b013e31816453a6.

Elberling, C., & Vejlby Hansen, K. V. (1999). Hearing instruments: Interaction with user preference. In: Rassmussen, A. N., Osterhammel, P. A., Andersen, T. et al. Auditory Models and Non-Linear Hearing Instruments, Proceedings of the 18th Danavox Symposium.

Ericsson, K. A., & Simon, H. A. (1980). Verbal reports as data. Psychological Review 87(3):215–251. doi:10.1037/0033-295X.87.3.215.

Ericsson, K. A., & Simon, H. A. (1984). Protocol analysis: Verbal reports as data. The MIT Press.

Ericsson, K. A., & Simon, H. A. (1993). Protocol analysis: Verbal reports as data (Revised edition). Revision Eds.: K. A. Ericsson and H. A. Simon. MIT Press.

Fox, M. C., Ericsson, K. A., & Best, R. (2011). Do Procedures for Verbal Reporting of Thinking Have to Be Reactive? A Meta-Analysis and Recommendations for Best Reporting Methods. Psychological Bulletin, 137(2), 316–344. 10.1037/a0021663.

Füllgrabe, C., & Moore, B. C. J.(2018)The association between the processing of binaural temporal-fine-structure information and audiometric threshold and age: A meta-analysis. Trends in Hearing 22: 2331216518797259. 10.1177/2331216518797259.

Gabrielsson, A. (1979). Dimension analyses of perceived sound quality of sound-reproducing systems. Scandinavian Journal of Psychology 20: 159–169.

Gatehouse, S., Elberling, C., & Naylor, G. (2006) Linear and nonlinear hearing aid fittings 2. Patterns of candidature. International Journal of Audiology 45(3): 153-71. 10.1080/14992020500429484.

Gδβwein, J. A., Rennies, J., Huber, R., Bruns, T., Hildebrandt, A., & Kollmeier, B.(2023). Evaluation of a semisupervised self-adjustment fine-tuning procedure for hearing aids. International Journal of Audiology 62(2): 159-171. 10.1080/14992027.2022.2028022.

Gδβwein, J. A., Rennies, J., Winneke, A., Hildebrandt, A., & Kollmeier, B.(2024). Evaluation of adjustment behaviour in a semi-supervised self-adjustment fine-tuning procedure for hearing aids. International Journal of Audiology, 63(5): 313–325. 10.1080/14992027.2023.2196601.

Güss, C. D.(2018). What is going through your mind? Thinking aloud as a method in cross-cultural psychology. Frontiers in Psychology, 9, 1292. 10.3389/fpsyg.2018.01292.

Hickson, L., Nickbakht, M., Timmer, B. H. B., & Dawes, P. (2024). Developing a prototype web-based decision aid for adults with hearing loss. International Journal of Audiology, 63(10), 819–826. 10.1080/14992027.2023.2279024.

Jensen, N. S., Hau, O., Nielsen, J. B. B., Nielsen, T. B., & Legarth, S. V. (2019). Perceptual Effects of Adjusting Hearing-Aid Gain by Means of a Machine Learning Approach Based on Individual User Preference. Trends in Hearing, 23: 1–23. 10.1177/2331216519847413.

Keidser, G., Brew, C., Brewer, S., Dillon, H., Grant, F., & Storey, L. (2005). The preferred response slopes and two-channel compression ratios in twenty listening conditions by hearing-impaired and normal-hearing listeners and their relationship to the acoustic input. International Journal of Audiology, 44(11): 656–670. 10.1080/14992020500266803.

Keidser, G., Dillon, H., Carter, L., & O’Brien, A. (2012). NAL-NL2 Empirical Adjustments. Trends in Amplification 16(4): 211-223. 10.1177/1084713812468511.

Kliesch, S., Chalupper, J., Lenarz, T., & Büchner, A.(2023). Evaluation of two self-fitting user interfaces for bimodal ci-recipients. Applied Sciences, 13(14), 8411. 10.3390/app13148411.

Kollmeier, B., & Kiessling, J. (2018). Functionality of hearing aids: state-of-the-art and future model-based solutions. International Journal of Audiology, 57(sup3), S3-S28. 10.1080/14992027.2016.1256504.

Kuk, F. K. (1999). How flow charts can help you troubleshoot hearing aid problems. The Hearing Journal, 52(10), 46–52. 10.1097/00025572-199910000-00005.

Kur§un, B., Shola, C., Cunio, I. E., Langley, L., & Shen, Y.(2025). Variability of preference-based adjustments on hearing aid frequency-gain response. Journal of Speech, Language, and Hearing Research, 68(4): 2006–2025. 10.1044/2024_JSLHR-24-00215.

Mackersie, C. L., Boothroyd, A., & Garudadri, H. (2020). Hearing aid self-adjustment: Effects of formal speech-perception test and noise. Trends in Hearing 24: 2331216520930545. 10.1177/2331216520930545.

MacPherson A., & Akeroyd, M. A. (2013). The Glasgow Monitoring of Uninterrupted Speech Task (GMUST): A naturalistic measure of speech intelligibility in noise. Proceedings of Meetings of Acoustics; 19(1): 050068. 10.1121/1.4799865.

Maidment, D. W., Coulson, N. S., Wharrad, H., Taylor, M., & Ferguson, M. A. (2020). The development of an mHealth educational intervention for first-time hearing aid users: combining theoretical and ecologically valid approaches. International Journal of Audiology, 59(7), 492–500. l0.1080/l4992027.2020.1755063.

Maykut, P., & Morehouse, R. (1994). Beginning Qualitative Research: A Philosophical and Practical Guide (1st ed.). Routledge. 10.4324/9780203485781.

McShefferty, D., Whitmer, W. M., & Akeroyd, M. A. (2016). The just meaningful difference in speech-to-noise ratio. Trends in Hearing 20: 2331216515626570. 10.1177/2331216515626570.

Nelson, P. B., Perry, T. T., Gregan, M., & van Tasell, D. J. (2018). Self-adjusted amplification parameters produce large between-subject variability and preserve speech intelligibility. Trends in Hearing, 22: 2331216518798264. 10.1177/2331216518798264.

Newell, A., & Simon, H. A. (1972). Human problem solving. Englewood Cliffs, New Jersey: Prentice-Hall.

Nielsen, J. (1992) The usability engineering life cycle. Computer 25(3): 12–22. 10.1109/2.121503.

Nisbett, R. E. & Wilson, T. D. (1977). Telling more than we can know: verbal reports on mental processes. Psychol Rev. 84(3):231–259. doi:10.1037/0033-295X.84.3.231.

Pedersen, T. H. (2015). [online] Perceptual characteristics of audio. DELTA, SenseLab. Available at: https://forcetechnology.com/-/media/force-technology-media/pdf-files/unnumbered/senselab/tech-document-perceptual-characteristics-of-audio-uk.pdf?la=en [Accessed 11/08/2025].

Perry, T. T., Nelson, P. B., & Van Tasell, D. J. (2019). Listener factors explain little variability in self-adjusted hearing aid gain. Trends in Hearing, 23: 2331216519837124. 10.1177/2331216519837124.

Pressley, M.; Afflerbach, P. Verbal Protocols of Reading; Routledge: New York, NY, USA, 1995.

Pryce, H., Burns-O’Connell, G., Smith, S., & Shaw, R. (2026) The lived experience of hearing loss: A systematic review with narrative synthesis. InternationalJournal ofAudiology 65(2): 131–144. 10.1080/14992027.2025.2523902.

Punch, J. L., & Robb, R. (1992). Prescriptive hearing aid fitting by parameter adjustment and selection. Journal of the American Academy of Audiology, 3(2), 94–100.

Rosenzweig, E.(2015). *Successful User Experience: Strategies and Roadmaps.*(1st ed.). Elsevier Science & Technology.

Sabin, A. T., Van Tasell, D. J., Rabinowitz, B., & Dhar, S. (2020). Validation of a self-fitting method for over-the-counter hearing aids. Trends in Hearing, 24: 2331216519900589. 10.1177/2331216519900589.

Saldana, J.(2016). The coding manual for qualitative researchers(3rd ed.). Thousand Oaks, CA: Sage.

Schweitzer, C., Mortz, M. S., & Vaughan, N. (1999). Perhaps not by prescription, but by perception. Hearing Review, 6(Suppl. 1): 58–62.

Skinner, B. F. (1977). Why I am not a cognitive psychologist. Behaviorism 5(2): 1–10. http://www.jstor.org/stable/27758892.

Stevens, S. S. (1959). Cross-modality validation of subjective scales for loudness, vibration, and electric shock. Journal of Experimental Psychology 57(4): 201–209. 10.1037/h0048957.

Vaisberg, J. M., Beaulac, S., Glista, D., Macpherson, E. A., Scollie, S. D. (2021) Perceived sound quality dimensions influencing frequency-gain shaping preferences for hearing aid-amplified speech and music. Trends in Hearing 25: 2331216521989900. 10.1177/2331216521989900.

Valentine, S., Dundas, J. A., & Fitz, K. (2011). Evidence for the use of a new patient-centered fitting tool. Hearing Review, 18(4): 28–34.

Van Someren, M. W., Barnard, Y. F., & Sandberg, J. A. C. (1994). The think aloud method: a practical approach to modelling cognitive processes. (Knowledge-based systems). Academic Press.

Watson, J. B. (1925). Behaviorism. New York: W. W. Norton.

White, B. (1974). [sound file] I Can’t Get Enough Of Your Love Babe. Can’t Get Enough, 20th century records.

Whitmer, W. M., Caswell-Midwinter, B., & Naylor, G. (2022) The effect of stimulus duration on preferences for gain adjustments when listening to speech. International Journal of Audiology, 61(11): 940–947. 10.1080/14992027.2021.1998676.

Whitty, J. A., Walker, R., Golenko, X., & Ratcliffe J. (2014). A think aloud study comparing the validity and acceptability of discrete choice and best worst scaling methods. PLoS One 9(4): e90635. DOI: 10.1371/journal.pone.0090635.

Wilson, T. D., & Schooler, J. W. (1991). Thinking too much: Introspection can reduce the quality of preferences and decisions. Journal of Personality and Social Psychology, 60(2): 181–192. DOI: 10.1037//0022-3514.60.2.181.

Wilson, T. D. (1994). The Proper Protocol: Validity and Completeness of Verbal Reports. Psychological Science, 5(5), 249–252. l0.111l/j.1467-9280.1994.tb00621.x.

Wundt, W. (1888). Selbstbeobachtung und innere Wahrnehmung [Self-observation and inner perception]. Philosophical Studies 1: 615–617.

Yang, Q., Hahn, S., Chang, B., van den Berg, A., & Olsen, G. (2017). Affordance of real-time personalization and adaptation of hearing aid settings. In: Stephanidis, C. (eds) HCI International 2017 - Posters’ Extended Abstracts. HCI 2017. Communications in Computer and Information Science, vol 714. Springer, Cham. 10.1007/978-3-319-58753-0_46.

